# Synergistic impact of three complement polymorphisms in the donor, not the recipient, on long-term kidney allograft survival

**DOI:** 10.1101/2023.10.24.23297481

**Authors:** Felix Poppelaars, Mariana Gaya da Costa, Bernardo Faria, Siawosh K. Eskandari, Vojtech Petr, V. Michael Holers, Mohamed R. Daha, Stefan P. Berger, Jeffrey Damman, Marc A. Seelen, Joshua M. Thurman

## Abstract

**Background:** Genetic analysis in transplantation offers potential for personalized medicine. Given the crucial role of the complement system in renal allograft injury, we investigated in kidney transplant pairs the impact of complement polymorphisms on long-term outcomes.

**Methods:** In this observational cohort study, we analyzed polymorphisms in C3 (*C3_R102G_*), factor B (*CFB_R32Q_*), and factor H (*CFH_V62I_*) genes of 1,271 donor-recipient kidney transplant pairs and assessed their association with 15-year death-censored allograft survival.

**Results:** Individually, only the presence of the *CFB_32Q_* variant in the donor and the combined presence in donor-recipient pairs were associated with better graft survival (*P*=0.027 and *P*=0.045, respectively). In the combined analysis, the *C3_R102G_*, *CFB_R32Q_*, and *CFH_V62I_* variants in the donor independently associated with the risk of graft loss (HR 1.32; 95%-CI, 1.08– 1.58; *P*=0.005). Thus, donor kidneys carrying the genetic variants that promote the highest complement activity exhibited the worst graft survival, whereas those with the genetic variants causing the lowest complement activity showed the best graft survival (15-year death-censored allograft survival: 48.8% vs 87.8%, *P*=0.001).

**Conclusion:** Our study demonstrates that the combination of complement polymorphisms in the donor strongly associates with long-term allograft survival following kidney transplantation. These findings hold significance for therapeutic strategies involving complement inhibition in kidney transplantation.

## 1. Introduction

The complement system can be activated through the classical, lectin, and/or alternative pathway, all leading to the formation of C3-convertases that cleave C3 into C3b and C3a.^1^ The newly generated C3b, in combination with factor B, can create more C3-convertases, resulting in an amplification loop with exponential potential.^2,3^ This amplification loop is tightly regulated by factor H, which rapidly dissociates factor B from the C3-convertase and catalyzes the inactivation of C3b through factor I-mediated cleavage to the iC3b form.^4^ Understandably, activation of the complement cascade is recognized as a critical proponent of kidney allograft injury throughout various phases of transplantation; in deceased donors before transplantation, during organ perfusion, following reperfusion, and in the recipient in various settings such as antibody-mediated rejection.^5–9^ Pre-clinical studies have shown promising results with complement inhibition in kidney transplantation to improve allograft outcomes.^10–14^ Furthermore, ongoing clinical trials with complement-targeting therapeutics aim to assess their impact on long-term outcomes after kidney transplantation.^7^

In recent decades, numerous studies have linked variations in complement genes to a wide range of inflammatory and infectious diseases.^15–17^ Functional characterization of these complement variants has yielded valuable insights into the underlying pathogenic mechanisms of complement activation in these disorders.^18^ Using *in vitro* experiments, recent studies revealed that individual polymorphisms only cause small changes in the activity of the complement system. However, when these complement variants are combined, their collective impact becomes significant.^19,20^ Thus, to achieve a comprehensive genetic understanding of the complement system, the total repertoire of polymorphisms in complement genes should therefore be studied, as this system operates as a network of proteins.^21^ The total make-up of the inherited set of complement variants is called the complotype and is believed to determine one’s individual ability to activate and regulate the complement system.^19,21^ A complotype leading to amplified complement activity makes an individual susceptible to inflammation, while a complotype that dampens complement activity increases the individual risks of infection.^20,21^

Several studies have demonstrated the impact of complement polymorphisms on outcomes after kidney transplantation.^22–27^ Yet, the majority of these studies focused on individual complement polymorphisms or haplotypes of a single complement gene. The combination of complement polymorphisms is likely to have a larger impact on long-term outcomes than one particular variant.^28^ Given the crucial role of complement in the pathogenesis of transplant renal injury, we, therefore, hypothesized that the complotype could be a major determinant of long-term allograft survival.^28^ Furthermore, genetic analysis in transplantation offers a unique opportunity to understand the effects of donor versus recipient genotype as well as develop personalized strategies that can improve donor-recipient matching, advance individualized risk stratification and drive personalized medicine.

As proof of concept, we examined the combined effect of three common complement polymorphisms (*C3_R102G_*, *CFB_R32Q_*, and *CFH_V62I_*) in donor-recipient pairs on long-term allograft survival after kidney transplantation. These three polymorphisms represent the most extensively researched and comprehensively characterized genetic variants within the complement system.^19,21^ The risk variant *C3_102G_*exhibits reduced binding to factor H compared with the reference *C3_102R_* variant, thereby amplifying complement activation.^19^ The protective *CFB_32Q_* variant, on the other hand, shows decreased potential to form C3- convertases and amplify complement activation compared to the reference *CFB_32R_* variant.^29^ Finally, the protective *CFH_62I_* variant demonstrates a higher capacity to bind C3b, catalyzing its inactivation and more effectively competing with factor B in C3-convertase formation compared to the reference *CFH_62V_* variant.^30^ The combination of these 3 genetic variants was selected based on prior research, which revealed that when the disease "risk" variants (*C3_102G_*, *CFB_32R_*, and *CFH_62V_*) were combined in a functional *in vitro* assay there was a sixfold higher complement activity compared than with the "protective" variants (*C3_102R_*, *CFB_32Q_*, and *CFH_62I_*).^19^ To address this hypothesis, we first analyzed the individual impact of complement variants in both the donor and recipient on 15-year death-censored allograft survival. Subsequently, we investigated the association of the combined presence of these variants with allograft survival.

## 2. Materials and methods

### 2.1 Study design

Patients receiving a first single kidney transplantation at the University Medical Center Groningen (UMCG) between March 1993 and February 2008 were selected for this study.^31–36^ Subsequent, exclusion criteria consisted of perioperative technical complications (3 pairs excluded), lack of available DNA (65 pairs excluded), loss of follow-up (4 pairs excluded), re-transplantation at the time of recruitment (22 pairs excluded) and simultaneous transplantation of other organs (65 pairs excluded). In summary, we began with 1430 kidney transplant pairs, and after applying exclusion criteria, we included 1271 kidney transplant pairs in this study. The endpoint was 15-year death-censored graft failure, defined as the return to dialysis or re-transplantation.

### 2.2 DNA extraction and genotyping

Peripheral blood mononuclear cells from blood or splenocytes were obtained from the kidney donors and their respective transplant recipients. DNA was isolated using commercial kits. DNA samples of 1,271 donor-recipient kidney transplant pairs was analyzed for three common complement polymorphisms, namely the *C3_R102G_* variant (rs2230199 C>G, Arg102Gly), the *CFH_V62I_* variant (rs800292 G>A, Val62Ile), and the *CFB_R32Q_*variant (rs641153 G>A, Arg32Gln). Genotyping of target single nucleotide polymorphism (SNP) was performed using the Illumina VeraCode GoldenGate Assay kit. Genotype clustering and calling were performed using BeadStudio Software. The overall genotype success rate was between 99.7 - 100%. Samples with a missing call rate were excluded from subsequent analyses, resulting in the exclusion of 12 donor-recipient pairs. Previously, we published the *C3* SNPs in this cohort.^26^ The distribution of all assessed SNP was in Hardy-Weinberg equilibrium.

### 2.3 Statistics

Statistical analysis was performed using SPSS 26.0. The data were presented as mean ± standard deviation (SD) for parametric variables, median [IQR] for non-parametric variables, and percentage [n (%)] for nominal data. To compare multiple groups, One-way ANOVA or Kruskal-Wallis *H-*test were used for normally and non-normally distributed variables, respectively, while χ2-tests were employed for categorical variables. The associations between complement polymorphisms and death-censored graft survival were assessed using Kaplan-Meier analyses with log-rank tests. Cox proportional-hazards regression analyses were performed to further examine the associations with graft loss, with adjustments made for potential confounders. Additionally, multivariable Cox regressions with stepwise forward selection were performed incorporating all variables that showed significant associations with graft loss in the univariable analyses. All statistical tests were two-tailed and *P*<0.05 was considered significant for all analyses.

### 2.4 Ethics

The study protocol was approved by the medical ethics committee at the UMCG under file number METc 2014/077 and the study was performed in accordance with the principles of the Declaration of Helsinki.

## 3. Results

### 3.1 The frequency of C3, CFH, and CFB polymorphisms in donor-recipient transplant pairs

We first examined the distribution of the *C3_R102G_, CFH_V62I_,* and *CFB_R32Q_* polymorphisms. There were no significant differences in the genotypic frequencies between the recipients and donors, also when compared to the European cohort of the 1000 Genomes Project (Table 1). Within the donor population, the number of minor alleles for the three complement polymorphisms per individual ranged from 0 to 4. Among the donors, 28.5% had no minor alleles, 41.9% had one, 22.5% had two, 6.6% had three, and 0.6% had four minor alleles (Figure 1). Similar results were seen in the recipient population; the number of minor alleles per individual ranged from 0 to 5, with 26.8% having no minor alleles, 43.1% having one, 22.6% having two, 6.6% having three, 0.5% having four, and 0.5% having five minor alleles (Figure 1). These results confirm that the occurrence of multiple complement polymorphisms within a single individual is frequently seen.

**Figure 1.**
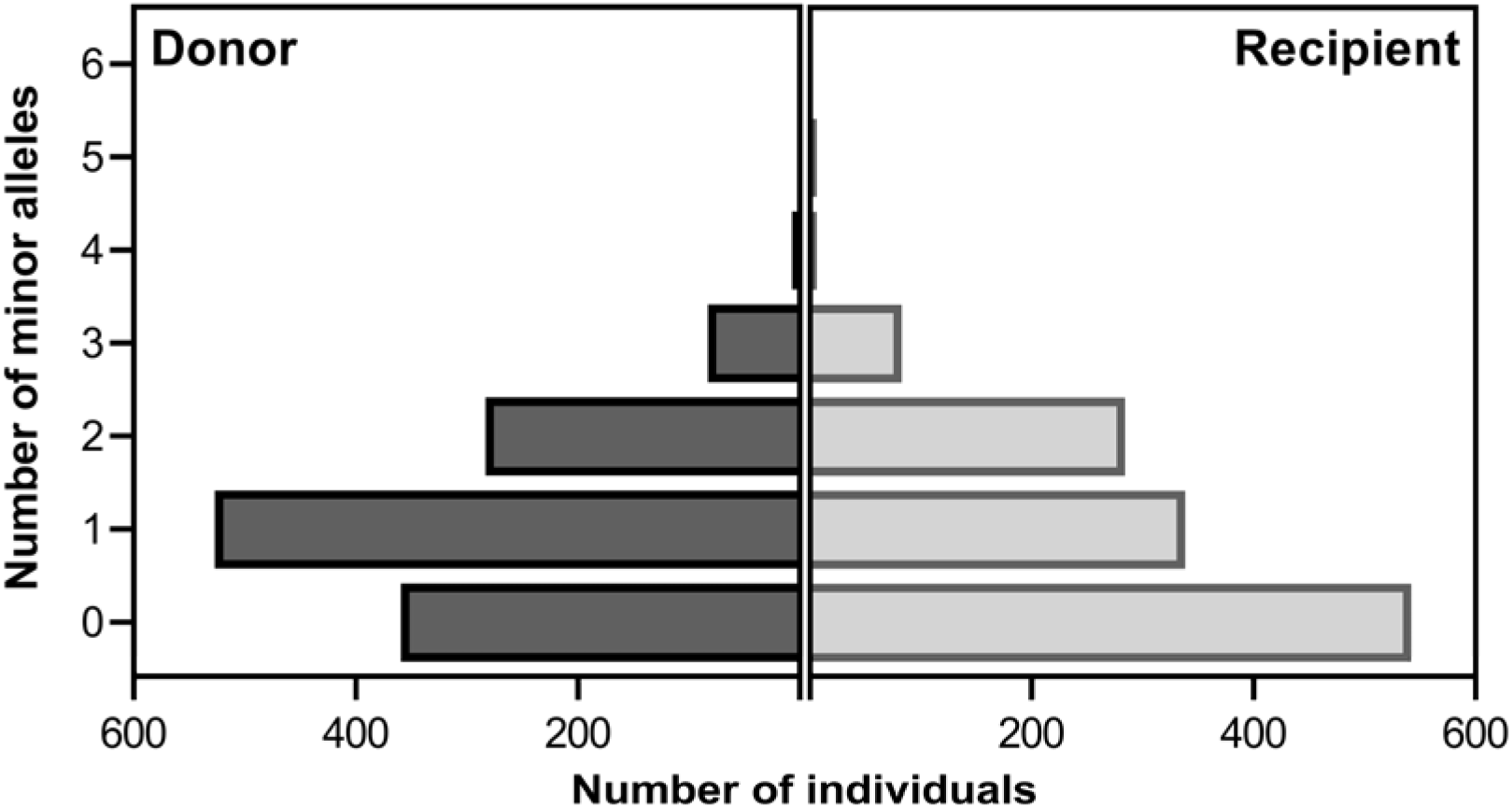
The number of minor alleles of complement polymorphisms in donors and recipients. The number of minor alleles of common polymorphic variants in *C3_R102G_* (rs2230199), *CFB_R32Q_* (rs641153), and *CFH_V62I_* (rs800292) was counted for each donor and recipient.

**Table 1:**
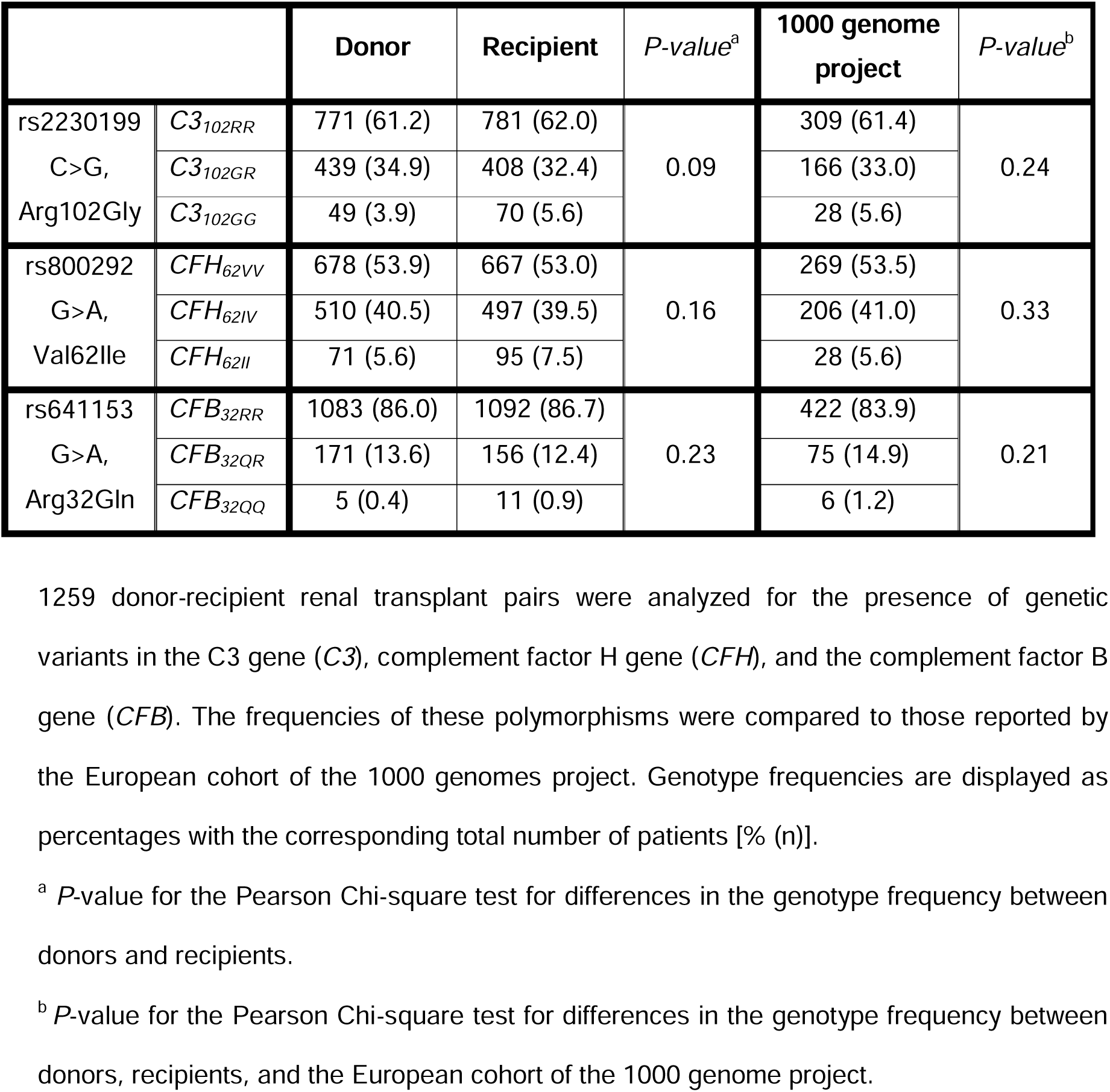
Genotypic frequencies of complement polymorphisms in donors and recipients.

|  |  | Donor | Recipient | *P*-value<sup>a</sup> | 1000 genome project | *P*-value<sup>b</sup> |
| --- | --- | --- | --- | --- | --- | --- |
| rs2230199<br>C>G,<br>Arg102Gly | *C3*<sub>102RR</sub> | 771 (61.2) | 781 (62.0) | 0.09 | 309 (61.4) | 0.24 |
|  | *C3*<sub>102GR</sub> | 439 (34.9) | 408 (32.4) |  | 166 (33.0) |  |
|  | *C3*<sub>102GG</sub> | 49 (3.9) | 70 (5.6) |  | 28 (5.6) |  |
| rs800292<br>G>A,<br>Val62Ile | *CFH*<sub>62VV</sub> | 678 (53.9) | 667 (53.0) | 0.16 | 269 (53.5) | 0.33 |
|  | *CFH*<sub>62IV</sub> | 510 (40.5) | 497 (39.5) |  | 206 (41.0) |  |
|  | *CFH*<sub>62II</sub> | 71 (5.6) | 95 (7.5) |  | 28 (5.6) |  |
| rs641153<br>G>A,<br>Arg32Gln | *CFB*<sub>32RR</sub> | 1083 (86.0) | 1092 (86.7) | 0.23 | 422 (83.9) | 0.21 |
|  | *CFB*<sub>32QR</sub> | 171 (13.6) | 156 (12.4) |  | 75 (14.9) |  |
|  | *CFB*<sub>32QQ</sub> | 5 (0.4) | 11 (0.9) |  | 6 (1.2) |  |

1259 donor-recipient renal transplant pairs were analyzed for the presence of genetic variants in the C3 gene (*C3*), complement factor H gene (*CFH*), and the complement factor B gene (*CFB*). The frequencies of these polymorphisms were compared to those reported by the European cohort of the 1000 genomes project. Genotype frequencies are displayed as percentages with the corresponding total number of patients [% (n)].
<sup>a</sup> *P*-value for the Pearson Chi-square test for differences in the genotype frequency between donors and recipients.
<sup>b</sup> *P*-value for the Pearson Chi-square test for differences in the genotype frequency between donors, recipients, and the European cohort of the 1000 genome project.

### 3.2 The association of single complement polymorphisms with allograft outcome

Next, we assessed the association between individual complement polymorphisms and late allograft loss. Kaplan-Meier analysis revealed significantly higher 15-year death-censored graft survival rate for donor kidneys carrying at least one ‘less activating’ *CFB_32Q_* variant when compared to donors who are homozygous for the reference *CFB*_32R_ variant (Fig. 2A, *P* = 0.027). After complete follow-up, graft loss incidence was 16.7% for renal grafts with at least one ‘less activating’ *CFB*_32Q_ variant and 28.1% for renal grafts carrying the reference *CFB*_32R_ variant. Conversely, no significant difference in long-term graft survival was observed based on the *C3_R102G_*and *CFH_V62I_*polymorphism in the donors (Supplementary Figure S1). Additionally, no significant difference in graft loss was seen for the *C3_R102G_, CFH_V62I_,* and *CFB_R32Q_* polymorphism in recipients (Supplementary Figure S2). In univariable analysis, the ‘less activating’ *CFB*_32Q_ variant in the donor was associated with a lower risk of graft loss and a hazard ratio of 0.60 (95%-CI, 0.38–0.95; *P* = 0.03). Subsequently, we performed a multivariable Cox regression analysis with a stepwise forward selection procedure using all variables that were significantly associated with graft loss in the univariable analysis (Table 2). In the final model, the ‘less activating’ *CFB_R32Q_*polymorphism in the donor, donor and recipient age, recipient blood type, and delayed graft function (DGF) were included. After adjustment, the ‘less activating’ *CFB*_32Q_ variant in the donor remained significantly associated with a lower risk of graft loss (HR, 0.55; 95%-CI, 0.34 – 0.89; *P* = 0.015). Overall, the presence of the *CFB*_32Q_ variant in the donor, which has reduced capacity to amplify complement activation, is linked to a lower risk of graft failure after kidney transplantation.

**Figure 2.**
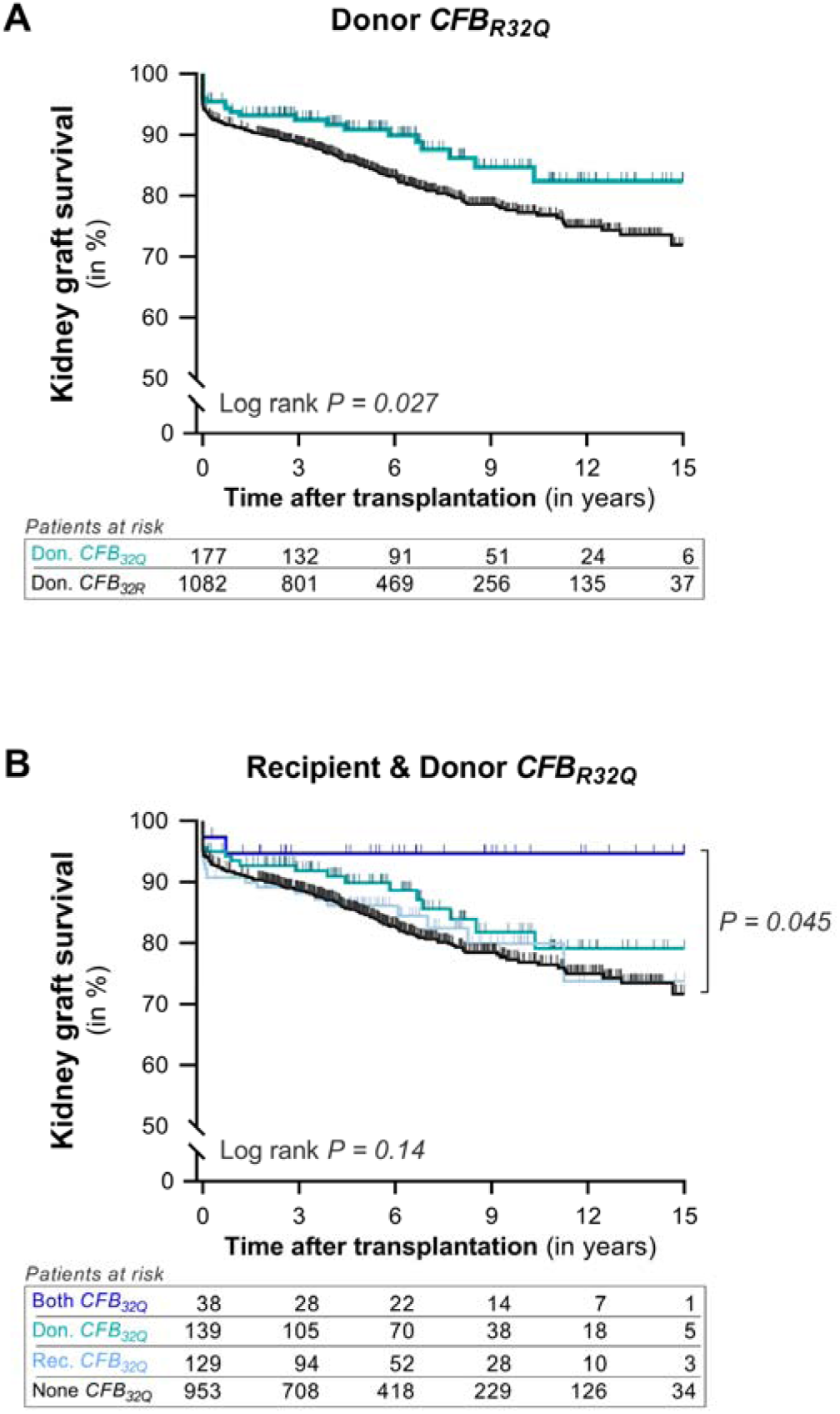
Kaplan–Meier curves of renal allograft survival according to the *CFB_R32Q_* variant. **(A)** Cumulative death-censored survival of kidney allografts based on the presence of the rs641153 complement factor B gene polymorphism (*CFB_R32Q_*) in allograft donors. Due to the low prevalence of donors homozygous for the *CFB_32Q_* variant, we merged heterozygote and homozygote donors into a single group. Donor kidneys carrying at least one *CFB_32Q_* variant (blue line – *CFB_32QR_+CFB_32QQ_*) were compared to donors who are homozygous for the reference *CFB*_32R_ variant (black line - *CFB_32RR_*). **(B)** Cumulative death-censored survival of kidney allografts based on the donor-recipient paired genotypes of the *CFB_32Q_* variant, comparing (i) pairs with both the donor and recipient being homozygous for the reference *CFB*_32R_ variant (black line), (ii) pairs where only the recipient carries the *CFB_32Q_* variant (light blue line), (iii) pairs where only the donor carries the *CFB_32Q_* variant (blue line), and (iv) pairs where both the donor and recipient carry the *CFB_32Q_* variant (dark blue line). Data represent death-censored survival curves and P-values were calculated using log-rank tests.

**Table 2:** Associations of the *CFB* polymorphism in the donor with graft loss.

| Variables | Hazard Ratio | P-value |
| --- | --- | --- |
| The *CFB*<sub>32R</sub> variant in the donor | 0.55 (0.34 – 0.89) | 0.015 |
| Delayed graft function (yes vs. no) | 4.09 (3.09 – 5.42) | <0.001 |
| Recipient age (in years) | 0.98 (0.97 – 0.99) | <0.001 |
| Recipient blood type (AB vs. other) |  | 0.002 |
| *AB* vs. *O* | 0.43 (0.26 – 0.71) | 0.001 |
| *AB* vs. *A* | 0.43 (0.26 – 0.70) | 0.001 |
| *AB* vs. *B* | 0.31 (0.15 – 0.61) | 0.001 |
| Donor age (in years) | 1.02 (1.01–1.03) | 0.003 |
| Warm ischemia time (in minutes) |  | 0.06 |
| Donor type (living vs. deceased) |  | 0.08 |
| Cold ischemia time (in hours) |  | 0.08 |
| Corticosteroids |  | 0.08 |
| Cyclosporin |  | 0.25 |
| Donor blood type (AB vs. other) |  | 0.94 |

Multivariable Cox proportional-hazards regression analyses for 15-year death-censored graft survival using a forward selection method. Variables with $P < 0.05$ in univariable analyses were included in the multivariable regression analyses. In the final model, the *CFB*<sub>R32Q</sub> polymorphisms in the donor, the occurrence of delayed graft function, recipient age, recipient blood type, and donor age were statistically significant, whereas donor transplant type, donor blood type, warm ischemia time, cold ischemia time, use of corticosteroids, and use of cyclosporin A were not. Data are presented as hazard ratios with 95% confidence intervals and $P$ -values of the Cox proportional-hazards analyses.
*CI*, confidence interval; *SD*, standard deviation; vs, versus.

Next, we explored whether the combined genotype of donor-recipient pairs for the *CFB_R32Q_* polymorphism exerted a more significant influence on graft survival than the donor genotype alone. Based on the *CFB*_32Q_ variant, donor-recipient pairs were divided into four groups (Fig. 2B). Recipients carrying the ‘less activating’ *CFB*_32Q_ variant that received a graft with the ‘less activating’ *CFB*_32Q_ variant demonstrated the most favorable outcome. Kaplan-Meier survival analyses revealed a significant difference in 15-year death-censored graft survival between donor-recipient pairs lacking the ‘less activating’ *CFB*_32Q_ variant and those where both the donor and recipient carried the ‘less activating’ *CFB*_32Q_ variant (graft loss: 28.3% versus 5.3%, *P* = 0.045). The ‘less activating’ *CFB*_32Q_ variant in the donor appeared to exert a more substantial impact on graft survival compared to its presence in the recipient (Fig. 2B). Collectively, these findings confirm that complement variants influence the risk of graft loss, and their functional consequence substantiates the role of complement in kidney transplant injury.

### 3.3 The association of the complotype with allograft outcome

Finally, we investigated the impact of the combined presence of the *C3_R102G_, CFH_V62I_,* and *CFB_R32Q_*polymorphisms on long-term allograft survival after kidney transplantation. Donor-recipient pairs were classified into 5 groups based on the combination of the three polymorphisms, referred to as the complotype. Analyses were performed separately for the complotype of the donor and the recipient. The groups consisted of the (i) high activity complotype, consisting of the gain-of-function *C3_102G_* variant and the reference variants for the other two polymorphisms (*C3_102G_/CFB_32R_/CFH_62V_*), the (ii) normal activity complotype, consisting of the reference variants for all three polymorphisms (*C3_102R_/CFB_32R_/CFH_62V_*), the (iii) dampened complotype, consisting of either the gain-of-function *CFH_62I_* or the loss-of-function *CFB_32Q_*variant with reference variants for the other two polymorphisms (*C3_102R_/CFB_32R_/CFH_62I_* or *C3_102R_/CFB_32Q_/CFH_62V_*), the (iv) mixed Complotype, consisting of the gain-of-function *C3_102G_* variant together with the gain-of-function *CFH_62I_* and/or the loss-of-function *CFB_32Q_*, and the (v) low activity complotype, consisting of both the gain-of-function *CFH_62I_*and the loss-of-function *CFB_32Q_* variant together with the reference *C3_102R_* variant (*C3_102R_/CFB_32Q_/CFH_62I_*).

Kaplan-Meier survival analysis revealed a significant difference in graft survival based on the donor’s complotype (Fig. 3; *P* = 0.024). Donor kidneys carrying the variant set that promotes the highest complement activity (high activity complotype – *C3_102G_/CFB_32R_/CFH_62V_*) had the shortest graft survival with a mean time to graft loss of 9.3 years, whereas donor kidneys with the variant set causing the lowest complement activity (low activity complotype – *C3_102R_/CFB_32Q_/CFH_62I_*) had the longest graft survival with a mean time to graft loss of 14.0 years (*P* = 0.001). After complete follow-up, the incidence of graft loss was 51.2% for donor kidneys with a high activity complotype, and 12.2% for donor kidneys with a low activity complotype. In univariable analysis, the donor’s complotype (as an ordinal variable) was significantly associated with 15-year death-censored graft survival (HR, 1.30; 95%-CI, 1.09 – 1.56; *P* = 0.005). When comparing donors with a low activity complotype to those with a high activity complotype, the HR for graft loss was 5.82 (95%-CI, 1.90 – 17.79; *P* = 0.002). In contrast, no association was observed between the recipient’s complotype and graft loss (Supplementary Figure S3, *P* = 0.99).

**Figure 3.**
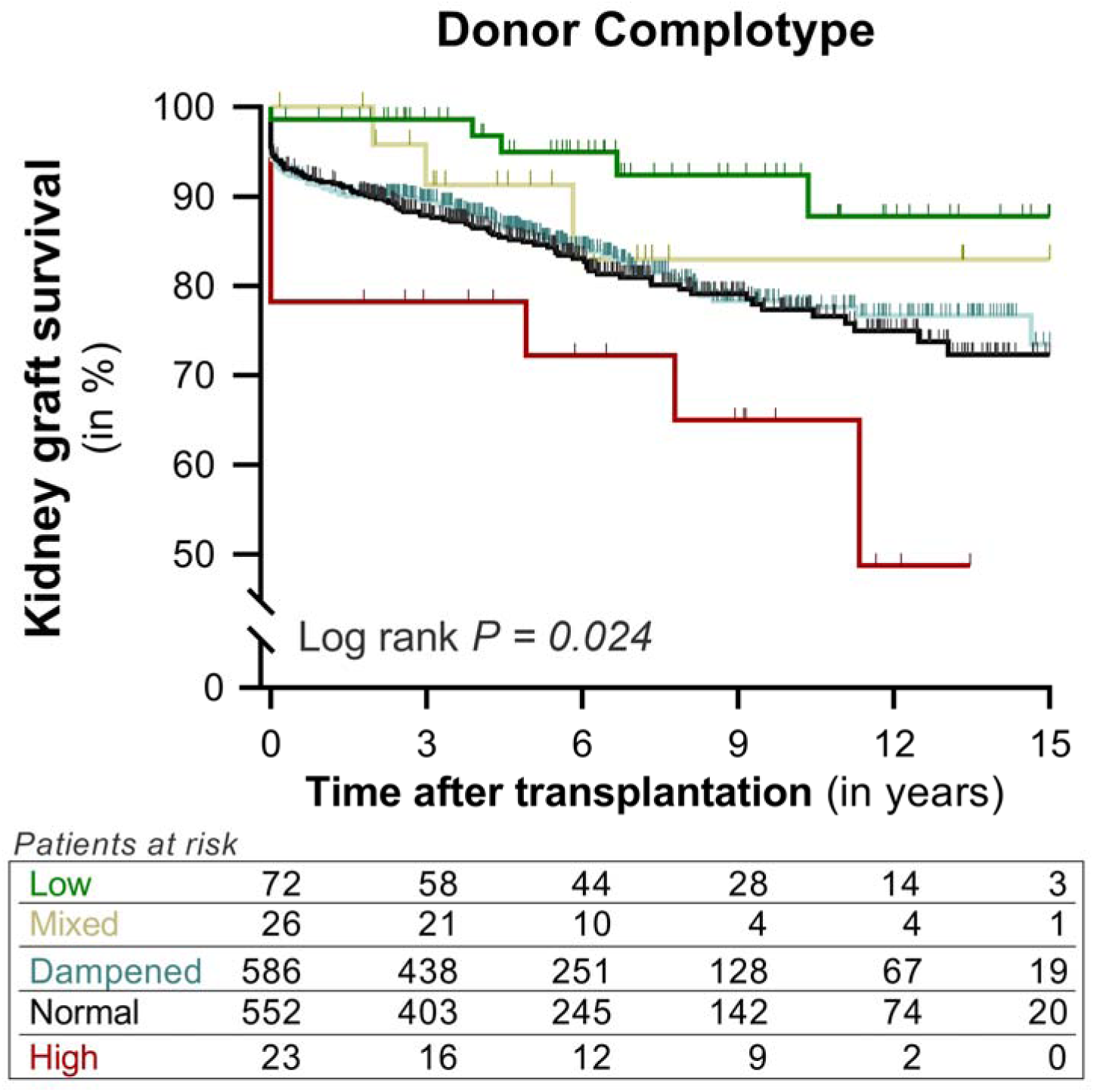
Kaplan–Meier curves of renal allograft survival according to the donor complotype. Cumulative death-censored graft survival of kidney transplants based on the combined presence of the *C3_R102G_, CFH_V62I_,* and *CFB_R32Q_* polymorphisms in allograft donors. The comparisons include: (i) high activity complotype (red line), which consists of the gain-of-function *C3_102G_*variant and the reference variants for the other two polymorphisms (*C3_102G_/CFB_32R_/CFH_62V_*); (ii) normal activity complotype (black line), consisting of the reference variants for all three polymorphisms (*C3_102R_/CFB_32R_/CFH_62V_*); (iii) dampened complotype (light blue line), consisting of either the gain-of-function *CFH_62I_* or the loss-of-function *CFB_32Q_* variant with reference variants for the other two polymorphisms (*C3_102R_/CFB_32R_/CFH_62I_* or *C3_102R_/CFB_32Q_/CFH_62V_*); (iv) mixed Complotype (yellow line), consisting of the gain-of-function *C3_102G_* variant together with the gain-of-function *CFH_62I_*and/or the loss-of-function *CFB_32Q_*; and (v) low activity complotype (green line), consisting of both the gain-of-function *CFH_62I_* and the loss-of-function *CFB_32Q_* variant together with the reference *C3_102R_* variant (*C3_102R_/CFB_32Q_/CFH_62I_*). The data are represented through death-censored survival curves, and p-values were calculated using log-rank tests.

We conducted further investigations to examine potential differences in clinical characteristics among donor-recipient pairs with different donor complotypes. The only significant variation observed was Sirolimus use among the groups (Table 3, *P* = 0.04). Nevertheless, when adjusting for Sirolimus use, the donor complotype remained significantly associated with graft survival in Cox regression analysis (HR, 1.30; 95%-CI, 1.08 – 1.56; *P* = 0.005). To ensure an accurate interpretation of death-censored graft survival, we also analyzed patient survival to exclude this as a potential confounding variable. All-cause mortality was comparable among the donor complotype groups (Table 3, *P* = 0.84). Multivariable analysis was then conducted with stepwise adjustments for relevant clinical variables (Table 4), including donor characteristics (model 2), recipient characteristics (model 3), and transplant variables (model 4). In these analyses, the donor complotype retained its significant association with graft loss, irrespective of adjusting for other clinical variables. Lastly, we performed a multivariate analysis with a stepwise forward selection procedure (Table 5). In the final model, donor complotype, donor and recipient age, recipient blood type, and DGF were included. Following adjustment, the donor complotype (as an ordinal variable) exhibited an association with graft loss, with a hazard ratio of 1.31 (95%-CI, 1.08 – 1.58; *P* = 0.005) using the low activity complotype as a reference. In summary, these analyses unveil the synergistic impact of the *C3*, *CFH*, and *CFB* polymorphisms in the donor on graft loss following kidney transplantation, and this association remains significant even when accounting for other determinants and potential confounders.

**Table 3:** Baseline characteristics of transplant pairs based on the donor complotype.

|  | **Low**<br>**Complotype**<br>(n = 72) | **Mixed**<br>**Complotype**<br>(n = 26) | **Dampened**<br>**Complotype**<br>(n = 586) | **Normal**<br>**Complotype**<br>(n = 552) | **High**<br>**Complotype**<br>(n = 23) | P-value |
| --- | --- | --- | --- | --- | --- | --- |
| **Donor** |  |  |  |  |  |  |
| Age, years | 42.4 ± 15.7 | 47.3 ± 11.3 | 44.4 ± 14.3 | 44.5 ± 14.6 | 41.4 ± 14.8 | 0.48 |
| Male sex, n (%) | 34 (47.2) | 14 (53.8) | 294 (50.2) | 281 (50.9) | 16 (69.6) | 0.43 |
| **Blood group** |  |  |  |  |  |  |
| Type O, n (%) | 27 (37.5) | 12 (46.2) | 289 (49.5) | 293 (53.2) | 13 (56.5) | 0.20 |
| Type A, n (%) | 37 (51.4) | 12 (46.2) | 227 (38.9) | 213 (38.7) | 9 (39.1) |  |
| Type B, n (%) | 8 (11.1) | 1 (3.8) | 55 (9.4) | 32 (5.8) | 1 (4.3) |  |
| Type AB, n (%) | 0 (0) | 1 (3.8) | 13 (2.2) | 13 (2.4) | 0 (0) |  |
| **Donor type** |  |  |  |  |  |  |
| Living, n (%) | 17 (23.6) | 6 (23.1) | 139 (23.7) | 113 (20.5) | 4 (17.4) | 0.64 |
| Brain death, n (%) | 47 (65.3) | 13 (50.0) | 353 (60.2) | 352 (63.8) | 15 (65.2) |  |
| Circulatory death, n (%) | 8 (11.1) | 7 (26.9) | 94 (16.0) | 87 (15.8) | 4 (17.4) |  |
| **Recipient** |  |  |  |  |  |  |
| Age, years | 47.1 ± 13.7 | 46.1 ± 12.9 | 47.5 ± 13.1 | 48.3 ± 13.9 | 51.4 ± 9.7 | 0.52 |
| Male sex, n (%) | 41 (56.9) | 16 (61.5) | 343 (58.5) | 318 (57.6) | 14 (60.9) | 0.99 |
| **Blood group** |  |  |  |  |  |  |
| Type O, n (%) | 23 (31.9) | 10 (38.5) | 251 (42.8) | 264 (47.8) | 11 (47.8) | 0.26 |
| Type A, n (%) | 39 (54.2) | 12 (46.2) | 248 (42.3) | 224 (40.6) | 9 (39.1) |  |
| Type B, n (%) | 9 (12.5) | 2 (7.7) | 58 (9.9) | 41 (7.4) | 3 (13.0) |  |
| Type AB, n (%) | 1 (1.4) | 2 (7.7) | 29 (4.9) | 23 (4.2) | 0 (0) |  |
| Dialysis vintage, weeks | 197 [85 – 266] | 174 [82 – 249] | 170 [86 – 269] | 172 [95 – 260] | 154 [94 – 298] | 0.93 |
| Highest PRA, in % | 15.0 ± 28.9 | 7.0 ± 19.9 | 8.7 ± 21.4 | 11.0 ± 24.6 | 13.6 ± 29.8 | 0.16 |
| **Induction immunosuppression** |  |  |  |  |  |  |
| Anti-CD3 Moab, n (%) | 0 (0) | 0 (0) | 9 (1.5) | 10 (1.8) | 0 (0) | 0.70 |
| ATG, n (%) | 7 (9.7) | 1 (3.8) | 41 (7.0) | 51 (9.2) | 2 (8.7) | 0.60 |
| Interleukin-2 RA, n (%) | 9 (12.5) | 5 (19.2) | 89 (15.2) | 92 (16.7) | 4 (17.4) | 0.86 |
| **Maintenance immunosuppression** |  |  |  |  |  |  |
| Azathioprine, n (%) | 7 (9.7) | 1 (3.8) | 34 (5.8) | 30 (5.4) | 0 (0) | 0.44 |
| Corticosteroids, n (%) | 69 (95.8) | 26 (100) | 550 (93.9) | 522 (94.6) | 22 (95.7) | 0.69 |
| Cyclosporin, n (%) | 62 (86.1) | 23 (88.5) | 497 (84.8) | 474 (85.9) | 19 (82.6) | 0.96 |
| Mycophenolic acid, n (%) | 50 (69.4) | 17 (65.4) | 418 (71.3) | 396 (71.7) | 15 (65.2) | 0.91 |
| Sirolimus, n (%) | 2 (2.8) | 3 (11.5) | 16 (2.7) | 14 (2.5) | 2 (8.7) | 0.044 |
| Tacrolimus, n (%) | 7 (9.7) | 3 (11.5) | 42 (7.2) | 40 (7.2) | 3 (13.0) | 0.69 |
| **Transplantation** |  |  |  |  |  |  |
| CIT, in hours | 16.7 [9.1 – 22.0] | 13.7 [7.5 – 19.5] | 17.5 [10.2 – 23.0] | 18.0 [11.4 – 23.3] | 16.3 [12.1 – 20.0] | 0.11 |
| WIT, in minutes | 38.0 [31.3 – 45.8] | 35.0 [30.0 – 45.0] | 37.0 [30.0 – 44.0] | 36.0 [31.0 – 45.0] | 40.0 [32.0 – 45.0] | 0.75 |
| Total HLA mismatches | 2 [1 – 3] | 2 [0 – 3] | 2 [1 – 3] | 2 [1 – 3] | 1 [0 – 3] | 0.30 |
| **Outcomes** |  |  |  |  |  |  |
| DGF, n (%) | 20 (27.8) | 12 (46.2) | 187 (31.9) | 186 (33.7) | 7 (30.4) | 0.49 |
| Mortality, n (%) | 10 (13.9) | 3 (11.5) | 87 (14.8) | 88 (15.9) | 2 (8.7) | 0.84 |

Data are displayed as mean ± standard deviation for parametric variables; median [IQR] for non-parametric variables, and nominal data as the total number of patients with the corresponding percentage [n (%)]. Bold values are used to show which testing was statistically significant ( $P$ -value < 0.05). $P$ -value indicates the $P$ -value for the differences in baseline characteristics between the groups, tested by one-way ANOVA or the Kruskal-Wallis test for continuous variables, with the $\chi^2$ test for categorical variables.
PRA, panel-reactive antibody; CD3, cluster of differentiation 3; ATG, Anti-thymocyte globulin; RA, receptor antagonist; CIT, cold ischemia time; WIT, warm ischemia time; HLA, human leukocyte antigen; DGF, delayed graft function.

**Table 4:**
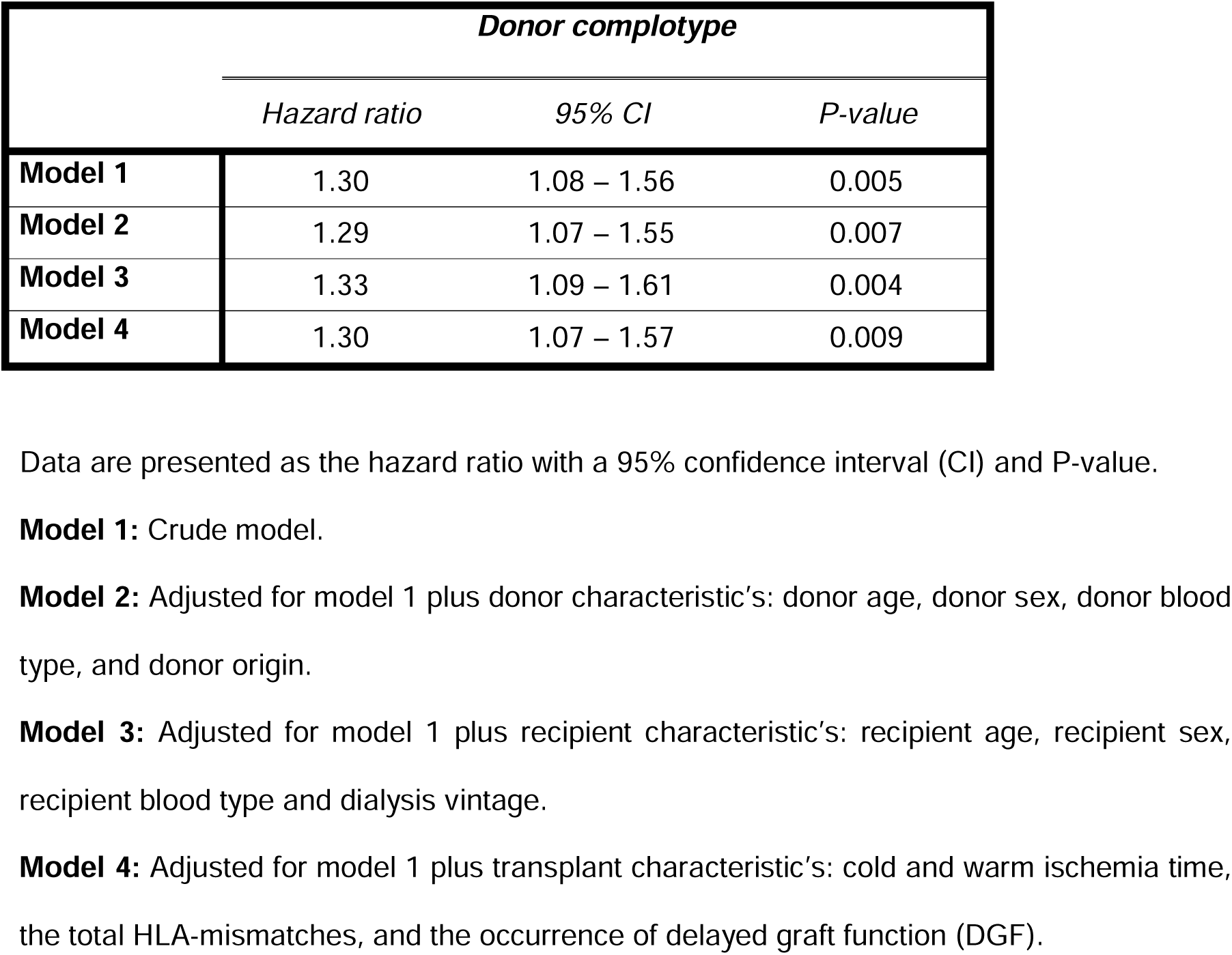
Associations of the complotype in the donor with graft loss.

| *Donor complotype* |  |  |  |
| --- | --- | --- | --- |
|  | *Hazard ratio* | *95% CI* | *P-value* |
| **Model 1** | 1.30 | 1.08 – 1.56 | 0.005 |
| **Model 2** | 1.29 | 1.07 – 1.55 | 0.007 |
| **Model 3** | 1.33 | 1.09 – 1.61 | 0.004 |
| **Model 4** | 1.30 | 1.07 – 1.57 | 0.009 |

Data are presented as the hazard ratio with a 95% confidence interval (CI) and P-value.
**Model 1:** Crude model.
**Model 2:** Adjusted for model 1 plus donor characteristic's: donor age, donor sex, donor blood type, and donor origin.
**Model 3:** Adjusted for model 1 plus recipient characteristic's: recipient age, recipient sex, recipient blood type and dialysis vintage.
**Model 4:** Adjusted for model 1 plus transplant characteristic's: cold and warm ischemia time, the total HLA-mismatches, and the occurrence of delayed graft function (DGF).

**Table 5:**
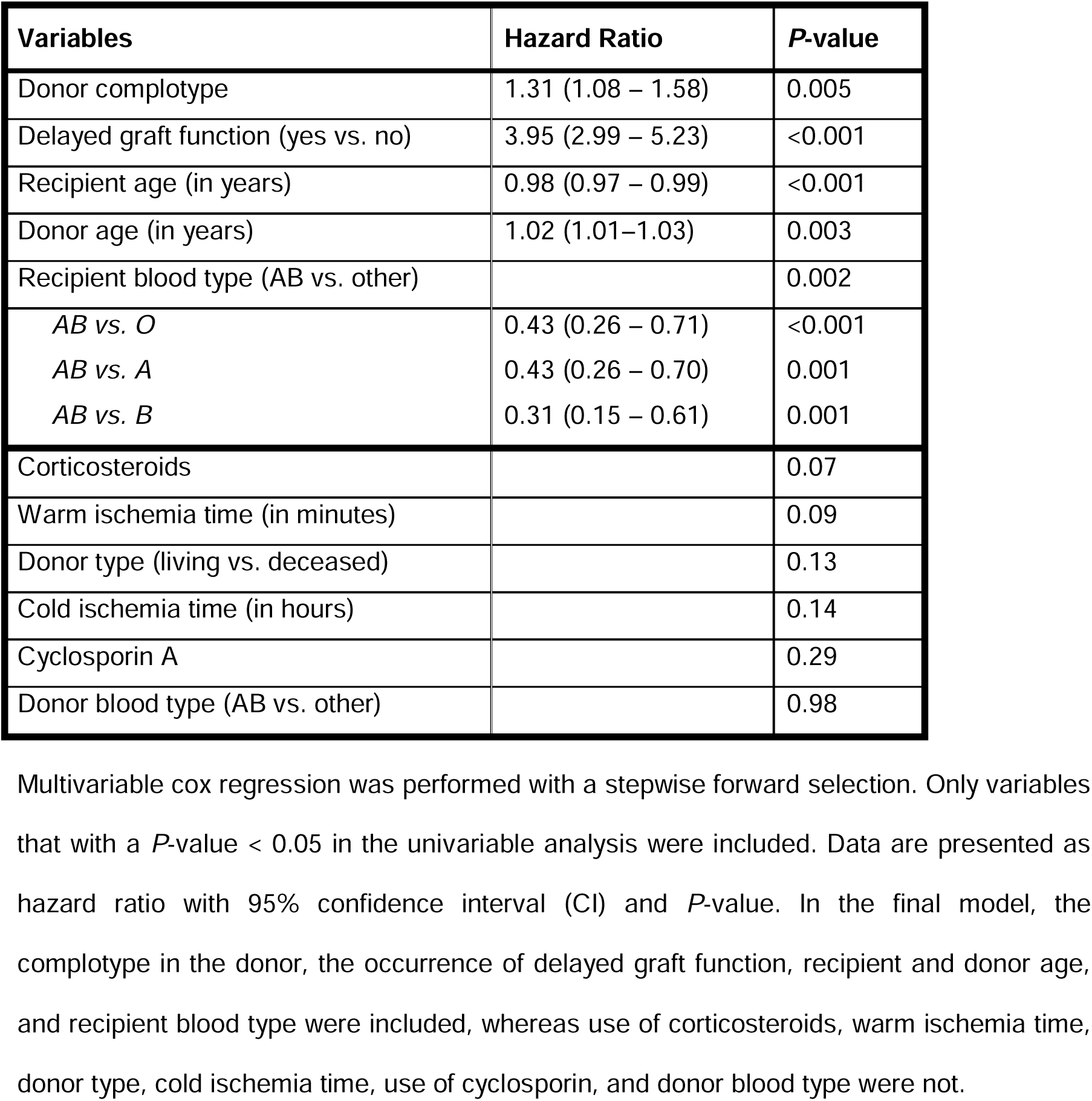
Competitive analysis of the associations of clinical factors with graft loss.

## 4. Discussion

Developing a comprehensive understanding of the immunological mechanisms underlying graft loss is crucial to create tailored treatment strategies in kidney transplantation.^37,38^ Studies of human genetics offer a complementary model to investigate mechanisms for the purpose of target validation.^39,40^ Moreover, therapeutic targets that are substantiated by human genetic evidence in disease association studies have a twofold higher probability of resulting in approved clinical drugs.^41,42^ The main finding of the current study is that disease-associated complement variants are common in both kidney transplant recipients and their donors, while these complement polymorphisms individually only have a modest impact on graft loss risk, when combined their collective effects significantly influence graft survival after kidney transplantation. Furthermore, our data support the notion that locally produced complement, originating from the donor kidney, plays a more prominent role in graft failure following kidney transplantation than complement generated by the recipient.

Evidence of the polymorphic nature of the complement system dates back to the 1960s, with the identification of two distinct C3 variants.^43^ At the protein level, the *C3_R102G_* polymorphism was classified as a slow variant (C3S – *C3_102R_*) and a fast variant (C3F – *C3_102G_*), based on the speed of C3 to migrate through a gel-electrophoresis system.^43^ Associations of the *C3_102G_* variant with different diseases has been well established, and the molecular basis for these disease associations was recently elucidated.^19^ Similarly, in the 1970s, the *CFB_R32Q_* variant was also first described at the protein level.^44^ Later on, it was shown that factor H was highly polymorphic as well.^45^ The relationship between the *C3_R102G_* polymorphism and outcome after kidney transplantation has been explored in multiple studies with varying results.^22,26,46,47^ To our knowledge, the current study is the first to examine the individual impact of the *CFB_R32Q_* and *CFH_V62I_*polymorphisms in both the donor and recipient on long-term graft survival after kidney transplantation. We observed a 34% risk reduction for graft loss in donor kidneys carrying at least one *CFB_32Q_* variant compared to kidneys homozygous for the reference *CFB_32R_* variant. Fittingly, the *CFB_32Q_* variant exhibits lower efficiency in amplifying complement activation. Notably, donor-recipient pairs both harboring the *CFB_32Q_* variant had the most favorable long-term outcome, revealing significant interactions between the complement systems of the donor and recipient, with a prominent role for donor-derived complement. In addition, our data strongly suggest that factor B presents an attractive therapeutic target in transplantation, further supported by preclinical models demonstrating the efficacy of factor B inhibitors in kidney transplantation.^11,48,49^ Altogether, these findings contribute to the growing body of evidence highlighting the pivotal role of the complement system in kidney transplant survival, underscoring the potential effectiveness of complement inhibitors to improve these outcomes.^5,6^

The concept of the complotype, defined as the complete inherited set of complement genes, was first introduced by Harris *et al*.^21^ Evidence supporting this concept was provided by *in vitro* experiments that simultaneously examined the combined effects of multiple polymorphic variants on complement activity. These studies revealed that individual complement gene variants had a relatively small impact, but when combined, their effects were additive and resulted in significantly higher complement turnover.^19,20^ Subsequently, *Paun et al*. provided the first *in vivo* evidence by showing an association between the combination of the *C3_102G_*, *CFB_L9H_*, *CFB_R32Q_*, and *CFH_V62I_* variants with higher systemic complement activation as well as with age-related macular degeneration (AMD).^50^ Presently, we found a dose-response relationship of a complotype composed of the *C3_R102G_*, *CFB_R32Q_*, and *CFH_V62I_*variants in the donor with long-term allograft survival. Furthermore, when comparing donor kidneys carrying the three "risk" variants (*C3_102G_/CFB_32R_/CFH_62V_*) to those carrying the three "protective" variants (*C3_102R_*/*CFB_32Q_*/*CFH_62I_*), the risk of graft loss was increased by approximately 400%. In contrast, the individual ’risk’ variants in the donor raised graft loss risk only by 51% for *CFB_32R_,* 33% for *C3_102G_*, and 17% for *CFH_62V_*. Mező *et al.* attempted to analyze the same combination in kidney transplantation, but their study focused solely on recipient genotypes, dividing them into two groups and assessing shorter graft survival. Nevertheless, graft survival was worse in recipients harboring at least one *C3_102G_*, *CFB_32R_*, and *CFH_62V_* variant.^51^ Another study demonstrated that the combined presence of two polymorphisms in the membrane-bound complement regulators CD46 and CD59 was associated with a favorable outcome after kidney transplantation, but the functional consequences of these variants are not well described, making their results harder to interpret.^27^ In addition to the additive effects of complement polymorphisms shown here, we have previously demonstrated the possibility of genetic compensation by different complement variants with opposing effects.^35^ Overall, these data present compelling evidence that instead of focusing solely on individual complement polymorphisms or complement genes for graft loss risk assessment, we should analyze the repertoire of common complement variants.

Clinical trials testing the efficacy of therapeutic complement inhibition in kidney transplantation have so far provided mixed results.^7^ Collectively, treatment of kidney transplant recipients with an anti-C5 monoclonal antibody did not result in major outcome improvements.^52,53^ Clinical trials involving C1-inhibitor in sensitized recipients yielded disappointing results as well.^54^ Yet, the perioperative administration of C1-inhibitor in kidney transplantation showed promising effects in a recent placebo-controlled randomized trial.^55^ The findings of our study offer valuable insights regarding the use of complement inhibitors in kidney transplantation, specifically on the site of action and patient selection. Importantly, we found that the complotype of the donor graft, rather than that of the recipient, was associated with the risk of graft loss. In accordance, Pratt *et al.* were to first to reveal the key role of donor-derived complement in allograft survival following experimental kidney transplantation in a murine model.^56^ Since then, accumulating data has further established the critical significance of locally produced complement in impacting outcomes after kidney transplantation.^26,57–62^ Moreover, the kidney significantly contributes to the circulating levels of complement.^63,64^ Altogether, this suggests that complement-targeted therapy in kidney transplantation should focus on the donor graft as the site of action. Immunosuppression in transplantation forms a delicate balance between the risks of graft rejection and infection, and while complement therapeutics may be beneficial in some patients, the harms may outweigh the benefits in others. Analyzing the complotype of donor-recipient pairs can identify those who are more susceptible to complement-mediated transplant kidney injury and may benefit from complement inhibition while avoiding over-immunosuppression in those with a low activity complotype, which would increase infectious risk without significantly improving graft outcomes. Further research is first needed to enable more comprehensive complotype testing that includes the analysis of functional polymorphisms of all complement genes. In addition, while the complotype sets an individual’s intrinsic complement activity, it does not provide information on activation. Ideally, the genetic analysis should thus be combined with additional tools such as plasma complement measurements and molecular imaging.^65,66^ Altogether, this could enable an individualized approach to help select individuals that would benefit from complement inhibition and help determine the timing, dosing, duration, and type of complement blocker needed.

This study has some limitations. While the association found in this study is expected to be causal, our study is prospective and observational in nature, which means we cannot definitively prove causality with our results. Additionally, due to the lack of available plasma samples, we could not measure complement activation in our cohort. However, it is worth noting that we deliberately selected the three most widely studied and functionally characterized complement polymorphisms, which helps strengthen the validity of our findings. The current study did not have sufficient statistical power to conduct donor-recipient complotype combinations or subgroup analyses (e.g., by sex, donor type, or recipient cause of renal failure) to explore the potential influence of these factors on the observed association. On the contrary, our study has several notable strengths, including the large size of the transplantation cohort, the combined analysis of the donor and recipient genotypes, the lengthy and thorough follow-up period, and the utilization of a robust and clinically significant endpoint (graft loss).

In conclusion, the current study has demonstrated that a complotype composed of *C3_R102G_*, *CFB_R32Q_*, and *CFH_V62I_* variants in the donor is strongly associated with long-term allograft survival following kidney transplantation. These findings are relevant in the context of therapeutic strategies for complement inhibition in kidney transplantation. The introduction of genomic and other molecular profiling techniques presents an unparalleled opportunity to implement precision pharmacotherapy in transplantation with the aim of enhancing patient outcomes. By utilizing genotype-based patient stratification, we can potentially identify donor-recipient pairs with a genetic predisposition to complement-mediated transplant kidney injury, rendering them excellent candidates for therapeutic complement inhibition in kidney transplantation. Furthermore, future studies should explore whether other complement polymorphisms in recipients can counteract the observed effects of the *C3_R102G_*, *CFB_R32Q_*, and *CFH_V62I_*variants in the donors.

## Abbreviations

AMD: Age-related macular degeneration
*C3*: Complement component 3 gene
*CFB*: Complement Factor B gene
*CFH*: Complement Factor H gene
DGF: Delayed graft function
HLA: Human leukocyte antigen
SNP: Single-nucleotide polymorphism

## Acknowledgements

The authors thank the members of the REGaTTA cohort (REnal GeneTics TrAnsplantation; University Medical Center Groningen, University of Groningen, Groningen, the Netherlands): S.J.L. Bakker, J. van den Born, M.H. de Borst, H. van Goor, J.L. Hillebrands, B.G. Hepkema, G.J. Navis and H. Snieder.

## Data availability statement

The data that support the findings of this study are available from the corresponding authors upon reasonable request.

## Author contributions

J.D. and M.A.S. formulated the overarching research aim and designed the study; F.P., and J.D. performed experiments and collected data; F.P. analyzed and interpreted the data; M.G.C., B.F., S.K.E., V.P., V.M.H., M.R.D., S.P.B., J.D., M.A.S., and J.M.T., critically reviewed the manuscript and provided commentary; F.P. wrote the first draft of the manuscript and revised it for publication; All authors read and approved the final manuscript.

