## Supplemental data for "Synergistic impact of three complement polymorphisms in the donor, not the recipient, on long-term kidney allograft survival"

**Supplemental Material**

**Figure S1**

**15-year death-censored kidney allografts survival stratified by the *C3_R102G_*, and *CFH_V62I_* variants in the donor**

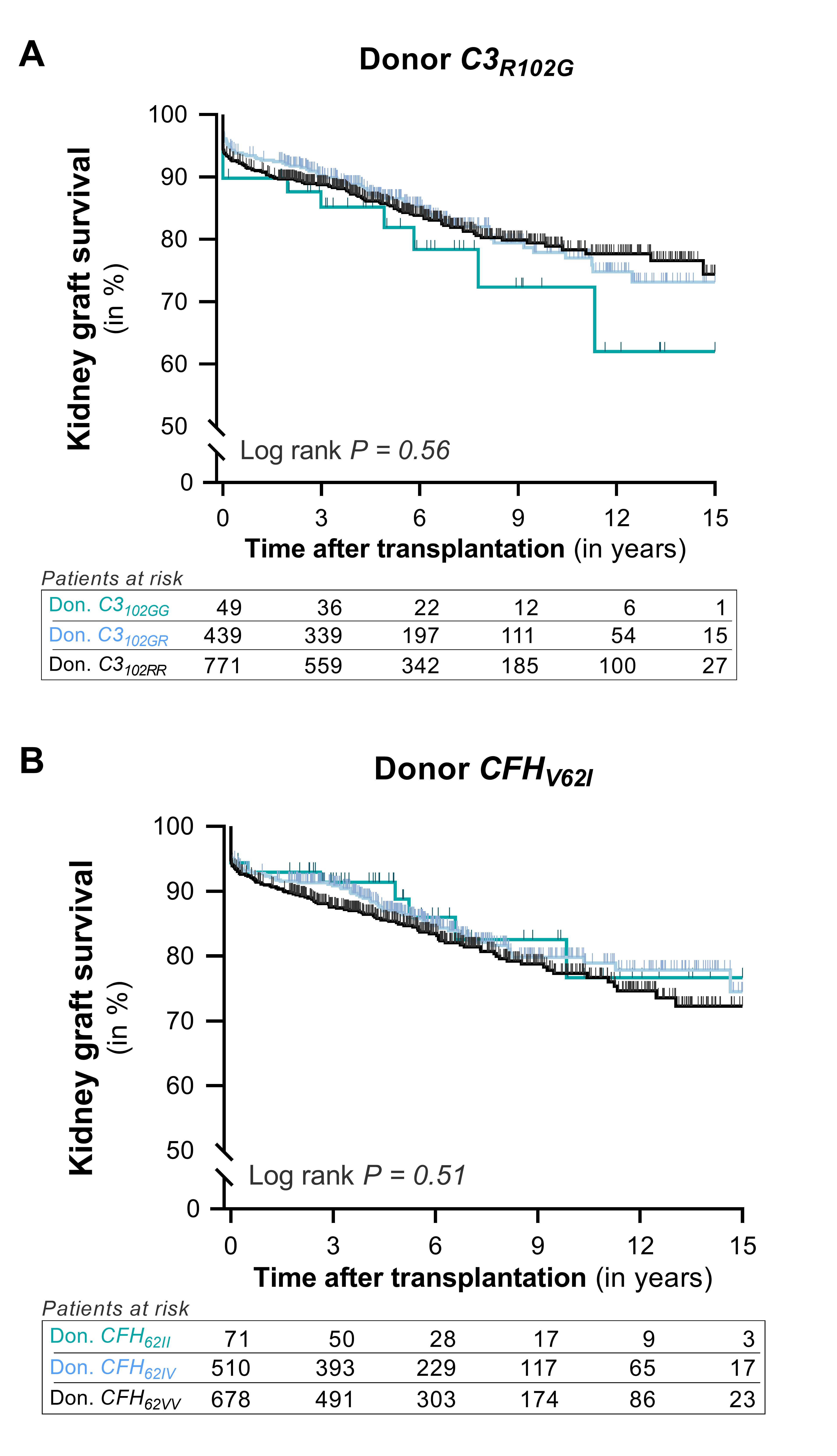

Cumulative death-censored survival of kidney allografts based on the presence of **(A)** the rs2230199 complement C3 gene polymorphism (*C3_R102G_*) and **(B)** the rs800292 complement factor H gene polymorphism (*CFH_V62I_*) in allograft donors. The data are represented through death-censored survival curves, and p-values were calculated using log-rank tests.

**Figure S2**

**15-year death-censored kidney allografts survival stratified by the *C3_R102G_*, *CFB_R32Q_* and *CFH_V62I_* variants in the recipient**

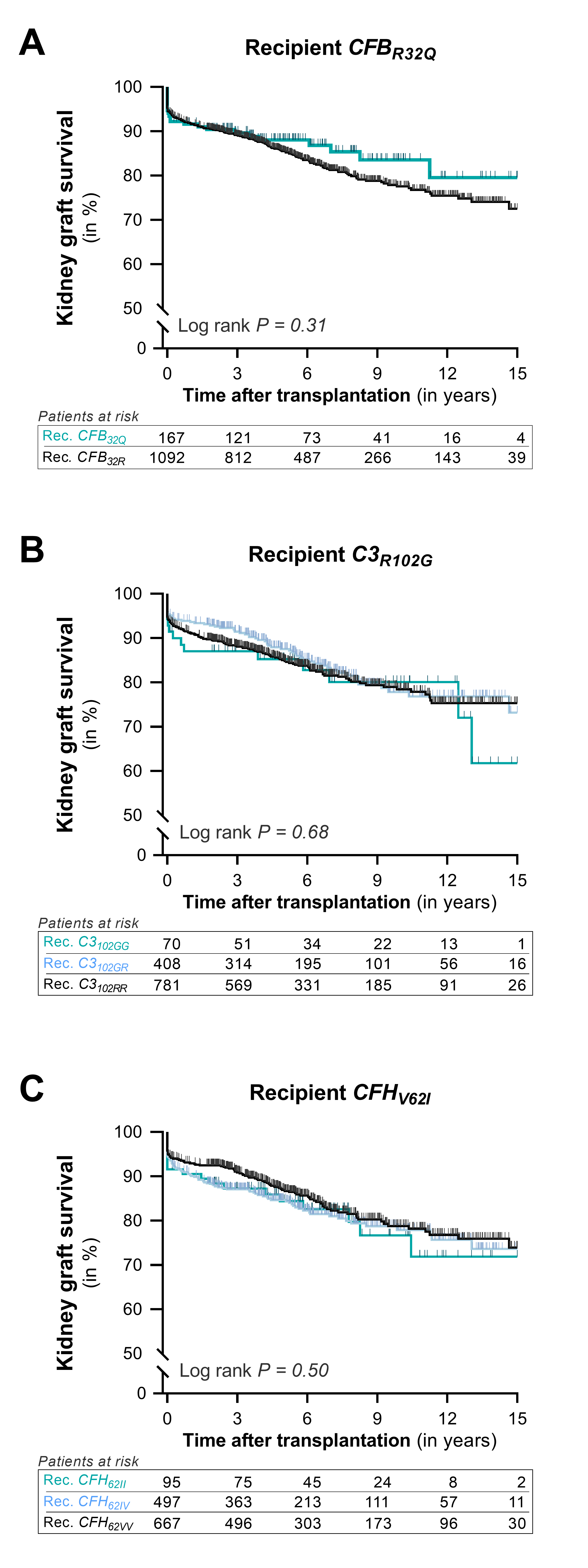

Cumulative death-censored survival of kidney allografts based on the presence of **(A)** the rs641153 complement factor B gene polymorphism (*CFB_R32Q_*), **(B)** the rs2230199 complement C3 gene polymorphism (*C3_R102G_*), and **(C)** the rs800292 complement factor H gene polymorphism (*CFH_V62I_*) in the recipient. The data are represented through death-censored survival curves, and p-values were calculated using log-rank tests.

**Figure S3**

**Kaplan–Meier curves of renal allograft survival according to the recipient complotype**

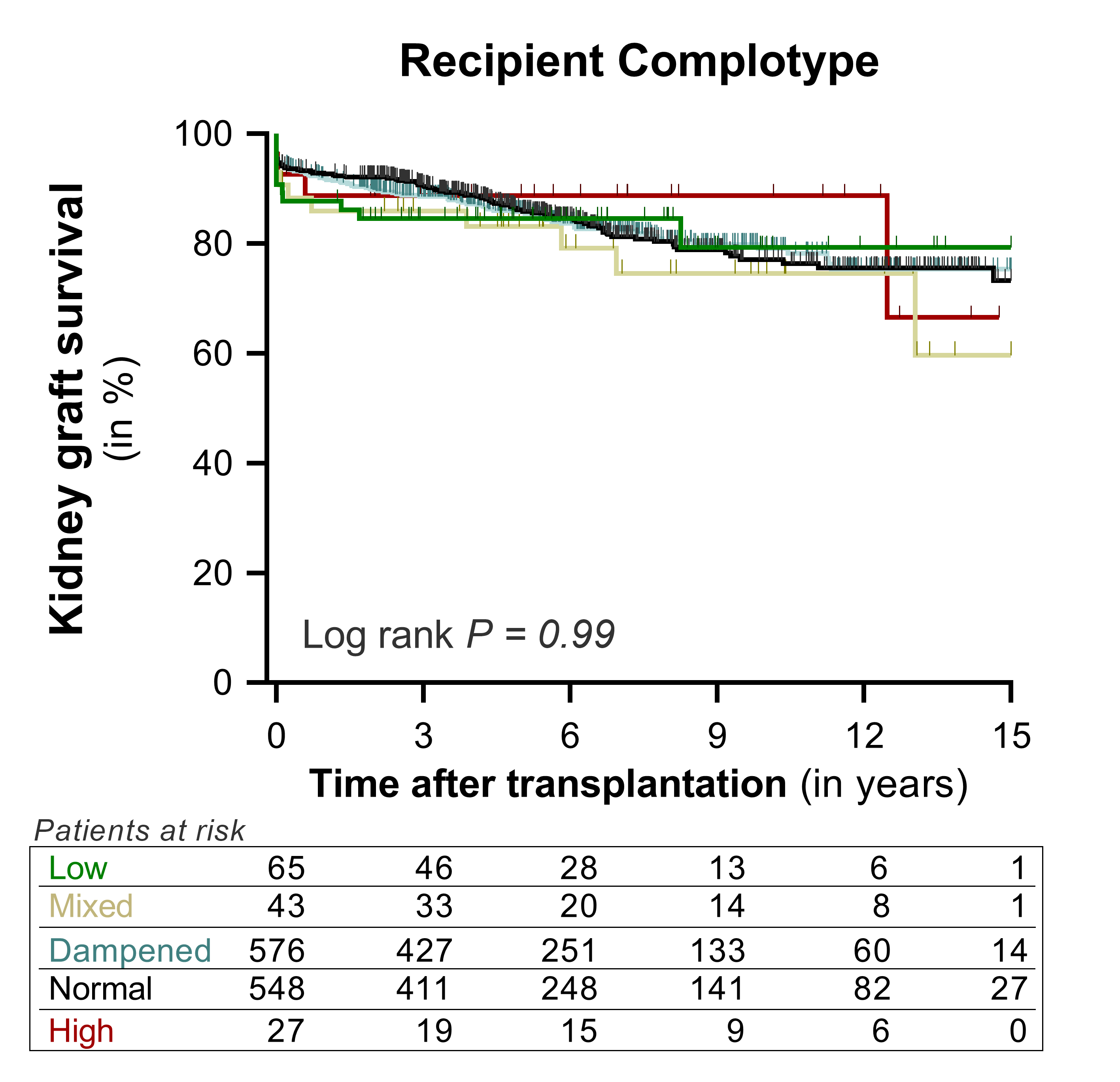

Cumulative death-censored graft survival of kidney allografts based on the combined presence of the *C3_R102G_, CFH_V62I_,* and *CFB_R32Q_* polymorphisms in kidney transplant recipients. The comparisons include: (i) high activity complotype (red line), which consists of the gain-of-function *C3_102G_* variant and the reference variants for the other two polymorphisms (*C3_102G_/CFB_32R_/CFH_62V_*)*;* (ii) normal activity complotype (black line), consisting of the reference variants for all three polymorphisms (*C3_102R_/CFB_32R_/CFH_62V_*); (iii) dampened complotype (light blue line), consisting of either the gain-of-function *CFH_62I_* or the loss-of-function *CFB_32Q_* variant with reference variants for the other two polymorphisms (*C3_102R_/CFB_32R_/CFH_62I_* or *C3_102R_/CFB_32Q_/CFH_62V_*); (iv) mixed Complotype (yellow line), consisting of the gain-of-function *C3_102G_* variant together with the gain-of-function *CFH_62I_* and/or the loss-of-function *CFB_32Q_*; and (v) low activity complotype (green line), consisting of both the gain-of-function *CFH_62I_* and the loss-of-function *CFB_32Q_* variant together with the reference *C3_102R_* variant (*C3_102R_/CFB_32Q_/CFH_62I_*). The data are represented through death-censored survival curves, and P-values were calculated using log-rank tests.
